# Structural and functional changes linked to cognitive impairment in Idiopathic Generalized Epilepsy

**DOI:** 10.64898/2026.03.11.26348164

**Authors:** Xinyuan Miao, Leo Chi U Seak, Wenying Du, Lingyuhao Zhang, Alisa Weng I Leong, Wenjing Yan, Yanping Sun

**Author notes:** Co-Correspondence author: Wenjing Yan; Department of Neurology, The Affiliated Hospital of Qingdao University, NO. 16 Jiangsu Road, Qingdao 266071, Shandong, China. Yanping Sun; Department of Neurology, The Affiliated Hospital of Qingdao University, NO. 16 Jiangsu Road, Qingdao 266071, Shandong, China.

## Abstract

**Background and Purpose:** While the idiopathic generalized epilepsy (IGE) comprise around one fifth of all epilepsy, the pathogenesis of it is largely unknown. Previous studies identified cognitive deficits in IGE patients, nevertheless, whether (and how) the brain structure and functional connectivity (FC) reflect these deficits remains underexplored. Here, we aim to find structural and FC differences in cognitively impaired IGE patients.

**Materials and Methods:** We recruited 36 IGE patients and 49 matched healthy controls (HC) in this cross-sectional study. All participants underwent structural and resting-state fMRI (rs-fMRI) scanning with a 3 Tesla MRI. Voxel-based morphometric analysis (VBM) was used to assessed structure differences, and seed-based analysis of rs-fMRI was used to examine FC. We examined the cognitive performance of patient with MoCA (Montreal Cognitive Assessment), grouped them into high (HMoCA, >25) and low (LMoCA, ≤25) group, and further examined the brain structural changes functional changes in each group.

**Results:** IGE patients showed right significant decrease in cerebellar gray matter volume (GMV), negatively correlating with the disease duration (r=-0.542, p=0.001), and increase in the left dorsolateral superior frontal gyrus GMV. Right cerebellum showed increased connectivity to the precuneus and angular gyrus, decreased connectivity to the postcentral gyrus and Rolandic operculum. Surprisingly, we found that LMoCA IGE patients (with more cognitive deficits) had increased right nucleus accumbens (NAc) GMV (t = -4.413, p < 0.001) and FC and a stronger NAc - prefrontal cortex FC (t = -2.683, p = 0.013), in comparison with the patients with high MoCA.

**Conclusions:** Cognitive impairment in IGE patients is linked to the NAc structural changes and NAc-prefrontal circuit alterations. These results provide novel circuit-level insights into understanding the cognitive impairment in IGE patients, contributing to revealing the pathophysiological mechanisms of IGE.

## Introduction

Idiopathic Generalised Epilepsy (IGE) is a neurological disorder that occurs predominantly in children and adolescents, accounting for nearly 20% of all epilepsy cases [1]. Patients with IGE usually exhibit generalized seizures (such as apoplectic seizures, myoclonic seizures, and tonic-clonic seizures), but lacking a prominent pathological sign [2, 3].

Despite the young age of IGE patients, cognitive dysfunction is surprisingly a common symptom in IGE patients, with studies reporting an impairment of attention, memory, and executive function in patients [4–7]. In other types of epilepsy, research has identified the dysfunction of different brain areas related to cognitive performance [8]. For example, Bell et al. demonstrated that cognitive challenges in memory, executive function, and language skills in the temporal lobe epilepsy patients are strongly linked to abnormalities in regions such as the hippocampus and frontal cortex, and their associated circuits [9, 10]. However, whether cognitive impairment in IGE is associated with functional and structural changes in our brain, have not been well investigated.

Clinical research in the last two decades has widely used Voxel-Based Morphometry (VBM) and Functional Connectivity (FC) to explore the abnormalities of cerebral structure and function in patients with neurological disorders [11, 12]. With these advances in neuroimaging, research has revealed that IGE, traditionally considered a structurally normal disease, exhibit subtle yet widespread changes in brain structure and function, offering new insights into its underlying mechanisms[13, 14]. For example, VBM studies have demonstrated distinctive reductions in grey matter volumes in the frontal, parietal, and temporal lobes of IGE patients. Similarly, FC analyses have identified abnormal connectivity in thalamo-cortical and fronto-thalamic circuits, as well as in large-scale resting-state networks including the default mode network (DMN) [15–18].

Nevertheless, these tools have not been used to study whether (and how) our brain changes relation to cognitive decline in IGE.

Nucleus accumbens (NAc) play an important role in multiple cognitive process including learning, value update, planning and cognitive control [19]. Lesions, atrophy, or FC dysfunction in NAc have found to be strongly associated the cognitive decline [20, 21]. In neurological diseases, such as Alzheimer’s and Parkinson’s diseases, researchers found significant reductions in volume in NAc, which is linked to the disease progression and cognitive function of the patients [20, 22–24]. Nevertheless, in epilepsy, NAc has not been well studied, and whether it may contribute to the cognitive function in epilepsy patients is unknown.

The present study aimed to systematically investigate the association of cognitive performance and the brain imaging features of IGE patients. As in previous epilepsy studies [25, 26], we adopted the Montreal Cognitive Assessment (MoCA) scale, and used it for the stratified fMRI analysis. We here found that the reduction nucleus accumbens volume and altered NAc - prefrontal cortex network is linked to the cognitive impairment in IGE patients, providing new insights into understanding the neural mechanisms of IGE.

## Methods

### Research Subject

Our study utilized a cross-sectional case-control design to collect demographic information and magnetic resonance imaging (MRI) data from IGE patients and healthy controls (HC). Both groups also thoroughly assessed clinical scales, ensuring a comprehensive analysis.

A total of 46 IGE patients and 49 healthy controls were recruited from the Department of Neurology at the Affiliated Hospital of Qingdao University between October 2021 and August 2024. Out of the 95 participants, two subjects with non-additive image quality and eight with excessive head motion were removed. A total of 85 subjects, including 36 patients and 49 HCs, met the quality control criteria and were included in the analysis. Inclusion criteria for IGE patients included (1) routine neurological examination; (2) normal background rhythm of the electroencephalogram (EEG); (3) an EEG showing all-conductance synchronous epileptiform discharges, which supported the diagnosis of generalized epilepsy; (4) no focal abnormalities on anatomical MRI. Patients on anti-seizure medications (ASMs) were instructed to withhold medication for 4 to 6 hours prior to the MRI scan. The diagnosis and subtyping of IGE were based on the criteria established by the International League Against Epilepsy (ILAE) [27]. Each patient was assigned to a specific IGE subtype—childhood absence epilepsy (CAE), juvenile absence epilepsy (JAE), juvenile myoclonic epilepsy (JME), or generalized tonic-clonic seizures alone (GTCSA)—according to clinical features (e.g., seizure semiology and age at onset), electroencephalographic (EEG) findings, and medical history. Two board-certified neurologists independently performed subtype classification, and discrepancies were resolved through discussion to reach a consensus.

HCs were recruited from the local community and matched to patients based on age and gender. They were all right-handed Han Chinese. Inclusion criteria for HCs were (1) the absence of systemic diseases and abnormal neurological signs, (2) the absence of a family history of neurological disorders, and (3) the absence of a history of convulsions or convulsive-like seizures. Exclusion criteria for all subjects included (a) neurological disorders other than IGE, (b) history of psychiatric disorders or medication for psychiatric disorders, (c) presence of contraindications to MRI, (d) excessive head movement during MRI imaging (average frame displacement exceeds 3 mm), (e) presence of comorbidities such as developmental delay or intellectual disability that could influence brain structure or cognition, and (f) exclusion of secondary generalized epilepsy (e.g., generalized epilepsy due to inflammatory or metabolic etiology).

All participants underwent the Montreal Cognitive Assessment (MoCA) test on the day of the MRI scan to assess their overall cognitive functioning. The participants also received the Hamilton Anxiety Scale (HAMA) and Hamilton Depression Scale (HAMD) tests to assess their anxiety and depression levels. Trained researchers conducted these evaluations under standardized conditions to ensure the consistency and reliability of the data.

We grouped the IGE patients based on their MoCA scores, categorizing those with scores ≤25 into the low MoCA score (LMoCA) group and those with scores >25 into the high MoCA score (HMoCA) group.

All research involving human participants was performed in accordance with the WMA Declaration of Helsinki. The study was approved by the Ethics Committee of Qingdao University Hospital, with informed consent obtained from all participants and/or their legal guardians before the study began.

### MRI Data Acquisition

All subjects underwent scanning using a Siemens MAGNETOM Skyra 3.0T MR scanner equipped with an 8-channel head coil for signal transmission and reception. Before entering the MR room, subjects were subjected to health screening and safety checks to rule out factors affecting image quality. During the scanning process, the subject lies flat on the examination bed, and foam cushions secure the sides of the head to minimize head movement. Subjects wore earplugs to minimize noise, stayed silent with their eyes closed, and avoided engaging in specific thoughts or falling asleep. All MRI scans were performed under standardized environmental conditions to ensure data accuracy and minimize interference from equipment or technical issues. The MRI protocol included: a. High-resolution T1-weighted (3D-T1) data were acquired using magnetization-prepared fast gradient echo sequences (TR=2530 ms, TE =2.27 ms, volume size:1mmx1mm x1mm matrix=256x256, layer thickness=1mm, FOV=256mm x256mm); b. Acquisition of resting-state functional magnetic resonance data using a planar echo imaging sequence (repetition time (TR)=2000 ms, echo time (TE)=30 ms, layer thickness =2.5mm, flip angle =90 °, matrix = 80x80, field of view (FOV) = 200 mm x 200 mm).

### Structural MRI Image Preprocessing

Structural image MRI data were preprocessed using SPM12 (https://www.fil.ion.ucl.ac.uk/spm) and CAT12 (https://neuro-jena.github.io/cat/) with T1 images normalized to template space. The brain images were then automatically segmented into grey matter (GM), white matter (WM), and cerebrospinal fluid (CSF). After preprocessing, the results of segmentation and normalization were assessed using the “Check data quality” tool of CAT12 by displaying slices of the samples and checking the sample homogeneity (including volume correlation analysis). Finally, Gaussian kernel smoothing (size 8mm full-width half height (FWHM)) was applied to the normalized grey matter images. After the above steps, the anomalous structure images were excluded.

### Functional MRI Image Preprocessing

The resting-state fMRI (rs-fMRI data) were preprocessed using SPM12 (https://www.fil.ion.ucl.ac.uk/spm) and DPABI V7.0 (https://rfmri.org/dpabi) with MatlabR2017b as the platform. Before preprocessing, the first 10 time points were removed to allow subjects to acclimatize to the MRI scanning environment and the signals to equilibrate. The remaining 198 volumes were preprocessed as follows: temporal layer correction; spatial normalization to standard Montreal Neurological Institute (MNI) space with 3 mm isotropic voxels; spatial smoothing using an isotropic Gaussian kernel with a 6 mm FWHM; removal of linear trends; temporal band-pass filtering (0.01 ∼ 0.08 Hz); and regression of interfering signals including the Friston 24 head motion parameters, global signal, white matter signal, and cerebrospinal fluid signal. Resting-state data exceeding 3 mm or 3 degrees of head movement in any direction were excluded.

Brain regions with significant differences in GMV were identified as masks, respectively. The masks were then set as regions of interest (ROIs) and seed regions. Whole-brain FC maps of ROIs were obtained using DPARSFA. Then, the time series between ROIs and other brain regions were extracted and averaged. Finally, the correlation coefficients were transformed into z-values using Fisher’s r-to-z transformation to improve the normality of the distribution.

## Data availability

The datasets used and/or analyzed during the current study are available from the corresponding author upon reasonable request.

## Statistical Analysis

Statistical analysis was conducted using the Statistical Package for the Social Sciences (SPSS)gr 20.0 software (Chicago, IL, USA). To compare the differences between the IGE group and HC regarding age, gender, MoCA, HAMA, and HAMD, the independent samples t-test was used for between-group comparisons; for categorical variables (e.g., gender, IGE subtypes, and seizure frequency), chi-square test were used. Besides, IGE patients were grouped based on MoCA scores, and subgroup analyses (LMoCA and HMoCA) were performed using independent samples t-test and chi-square test for all the two groups’ demographic and clinical scale scores.

Differences in GMV profiles between the IGE/HCs and HMoCA/LMoCA groups were calculated separately using independent samples t-tests. Age, sex, total intracranial volume (TIV), and years of education were included as covariates to control for potential effects. Multiple comparisons were performed using Gaussian random field correction (GRF correction) with clustering thresholds set at voxel *p* < 0.005 and cluster *p* < 0.05.

Clusters with significant differences in GMV between the IGE/HCs and HMoCA/LMoCA groups were selected as ROIs for FC analyses. Differences between the two groups were analyzed using voxel-based independent samples t-tests with age, sex, years of education, and TIV as covariates, and significantly different clusters were used as masks. Multiple comparisons were corrected using the GRF correction. These FC analyses set thresholds at voxel *p* < 0.001 and cluster *p* < 0.05.

Subgroup analyses were also performed for two major IGE subtypes: Juvenile Myoclonic Epilepsy (JME) and Generalized Tonic-Clonic Seizures on Awakening (GTCSA). The same statistical procedures as those used in the MoCA-based subgroup analysis were applied, including covariates (age, sex, TIV, and years of education). The same statistical thresholds as in the MoCA-based subgroup analysis were used. For VBM analyses, multiple comparison correction was performed using GRF correction with a voxel-level threshold of p < 0.005 and a cluster-level threshold of p < 0.05. For FC analyses, GRF correction was applied with a more stringent voxel-level threshold of p < 0.001 and cluster-level threshold of p < 0.05. Detailed results and relevant statistical information are provided in the Supplementary Material.

Pearson correlation analysis was conducted to examine the relationships between GMV, FC, and clinical variables, including disease duration, MoCA scores, HAMA, and HAMD. The analysis assessed the linear correlations between these variables, with correlation coefficients (r) and corresponding p-values reported. A two-tailed test was used, and statistical significance was set at *p* < 0.05.

This study employed linear regression to examine the effects of GMV and FC in abnormal brain regions on MoCA scores. Separate regression analyses were conducted to assess the individual impact of GMV and FC, controlling for age, gender, and education. Multifactorial linear regression analyses were then performed to account for potential confounding factors, with statistical significance set at *p* < 0.05. Additionally, sensitivity analyses were conducted by including medication use, seizure frequency, and disease duration as additional covariates. The statistical procedures were then repeated to evaluate the robustness of the results. To examine the effect of clinical heterogeneity on cognitive performance and radiographic features, we performed multiple linear regression analyses with radiographic measures as dependent variables. Age, gender, years of education, TIV, number of antiepileptic drugs, disease duration, and seizure frequency were used as covariates to control for confounding factors. Assumptions of normality, linearity, and homoscedasticity were verified, and multicollinearity between variables was assessed.

## Results

### Demographic and Clinical Characteristics

We included a total of 36 patients (19 women, with mean± SD age 22.83± 8.57 years) and 49 healthy controls (29 women, with mean± SD age 24.57± 2.27 years) in this study. In terms of age and gender, there were no significant (p>0.05, independent-samples t-test and chi-square test) differences between the IGE group and the HC group and between the HMoCA group and the LMoCA group. Consistent with previous research[28, 29], the MoCA scores of the IGE group were lower than those of the HC group (t=-10.20, p<0.001), while the HAMD (t=5.49, p<0.001) and HAMA (t=4.59, p<0.001) scores were higher than those of the HC group. There were more years of education in the HC group than in the IGE group (t=3.84, p<0.001), and there was no significant difference in the years of education between the HMoCA group and the LMoCA group. For patients with IGE, there were significant differences in MoCA scores (t=7.43, p<0.001), disease duration (t=2.72, p=0.010), and seizure frequency (^2^=4.01, p=0.045) between the HMoCA and LMoCA groups. However, no significant differences were found in the number of medications, IGE subtype, or HAMA and HAMD scores in the subgroup analyses (see Table 1).

**Table 1.**
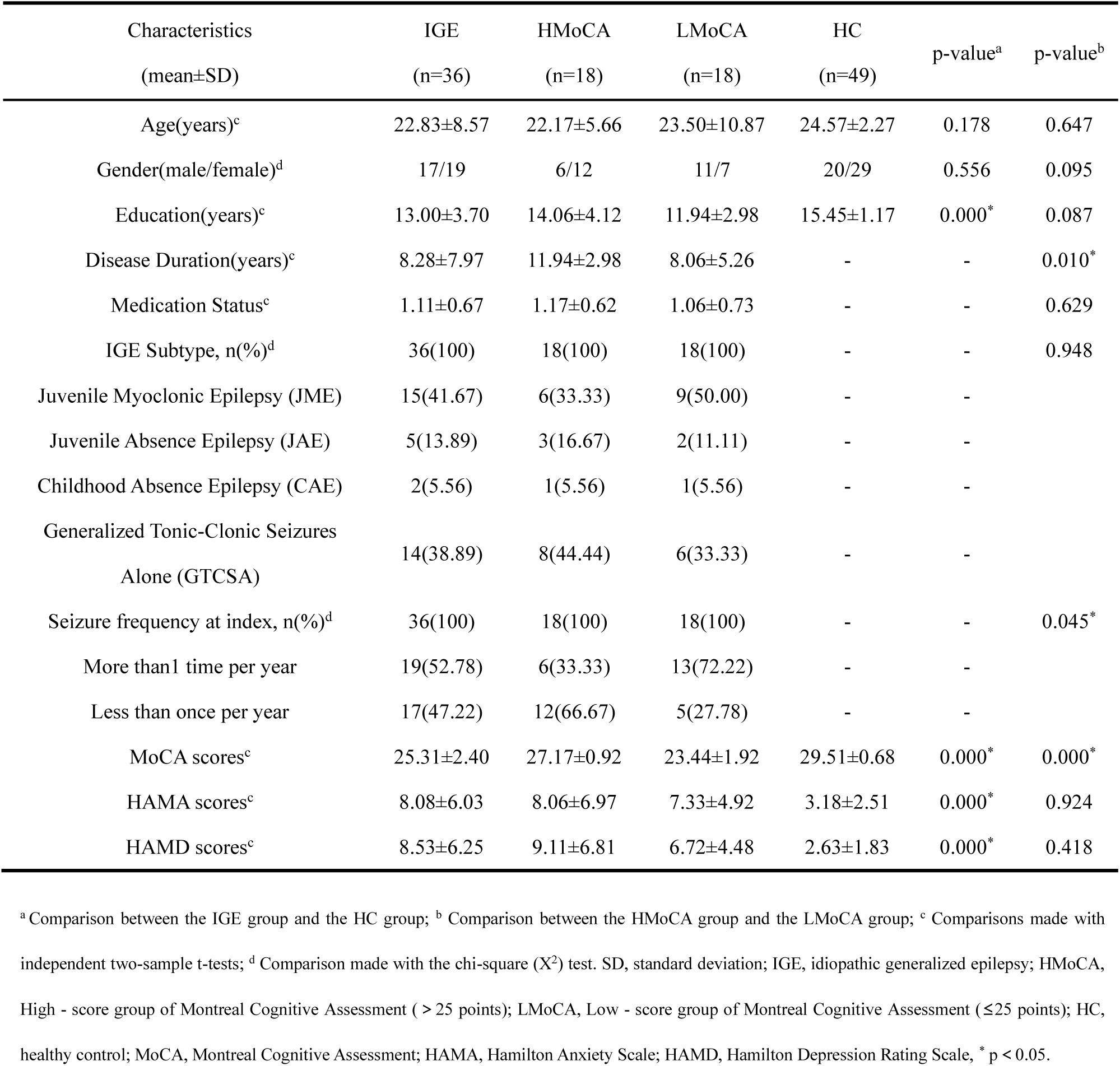
Baseline demographic and clinical characteristics.

### Brain structural changes in IGE patients relate to cognitive impairment

We next tested if there are differences in the brain structure between groups. Consistent with previous IGE studies [30–32], we found that the Gray Matter Volume (GMV) was significantly reduced in the right cerebellum (Cerebelum_8_R) and increased in the left dorsolateral superior frontal gyrus (Frontal_Sup_2_L) in the IGE group compared with the HC group.

We then examined if there are structural differences between IGE patients with more cognitive deficits (HMoCA) and those with less cognitive deficits (LMoCA). Intriguingly, we found significant GMV increase in the right nucleus accumbens (N_Acc_R) in the LMoCA group compared to the HMoCA group. (see Table 2, Fig. 1; GRF corrected, cluster level p < 0.05 with voxel-level threshold p < 0.005).

**Fig. 1.**
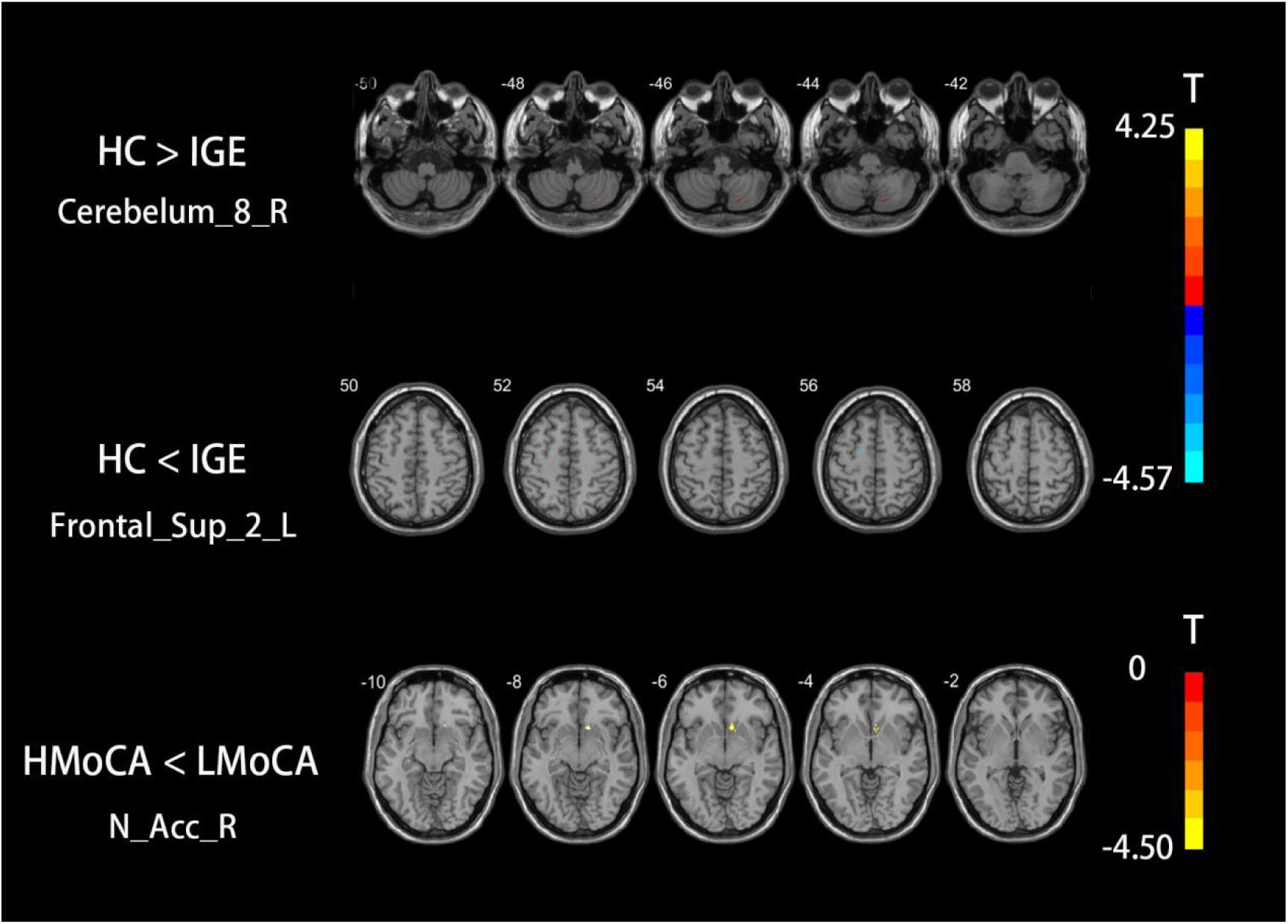
Voxel-based morphometry analysis results between groups. The color bar represents the t-value. Warm colors indicate the brain regions with reduced gray matter volume in IGE patients and HMoCA group, and cool colors indicate the brain regions with increased gray matter volume in IGE patients. The map threshold was set at p < 0.05 and was corrected by GRF.

**Table 2.** Results of VBM analyses.

| Groups | Cluster regions(AAL) | MNI coordinates | Cluster size<br>(Voxels) | Peak t value | p value |
| --- | --- | --- | --- | --- | --- |
|  |  | X, Y, Z |  |  |  |
| HC > IGE | Cerebelum_8_R(108) | 21.25 -73.25 -46.25 | 78 | 4.25 | 0.00** |
| HC < IGE | Frontal_Sup_2_L(3) | -19.75 -2.25 55.75 | 77 | -4.57 | 0.00** |
| HMoCA < LMoCA | N_Acc_R(158) | 6.25 13.75 -5.25 | 178 | -4.50 | 0.00** |

AAL, Anatomical Automatic Labeling; MNI, Montreal Neurological Institute; IGE, idiopathic generalized epilepsy; HC, healthy control; L, left; R, right.
\* p < 0.05; \*\* p < 0.01.

### Disrupted functional connectivity (FC) in IGE linked to cognitive decline

Compared with HCs, IGE patients had increased FC between the right cerebellum and specific brain regions (bilateral precuneus, right frontal_Sup_2_R, right horn R, and left cerebellar Crus2_L) but decreased FC (GRF-corrected) with the right posterior medial cortex and right Rolandic_Oper_R (Table 3, Fig. 2). In addition, compared with HCs, patients with IGE showed increased FC between the left frontal lobe and multiple other regions, including the Frontal_Sup_Medial_L, Temporal_Mid_L, Rectus_L, Lingual_L, Occipital_Sup_R, Temporal_Inf_L, and Temporal_Mid_R (Table 4, Fig. 3). Furthermore, patients in the LMoCA group had increased FC (GRF-corrected) between the right NAc and Frontal_Sup_2_R compared with the HMoCA group (Table 5, Fig. 4).

**Fig. 2.**
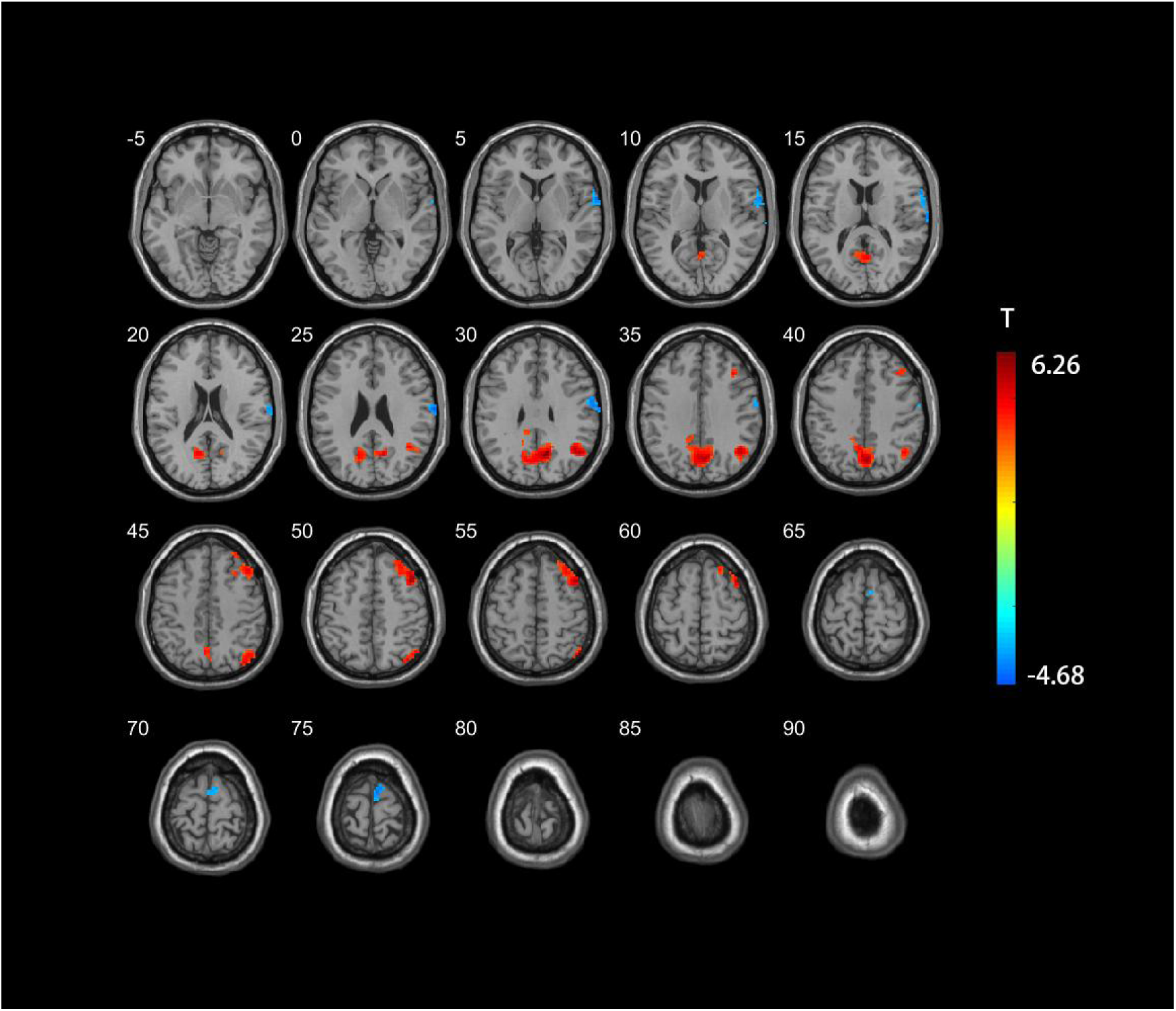
Altered functional connectivity from the right cerebellum to other brain regions in IGE patients. The color bar represents the t-value. Warm colors indicate increased and cool colors indicate decreased functional connectivity from the right cerebellum in IGE patients. The map threshold was set at voxel-level p < 0.001 and cluster-level p < 0.05, corrected by GRF.

**Fig. 3.**
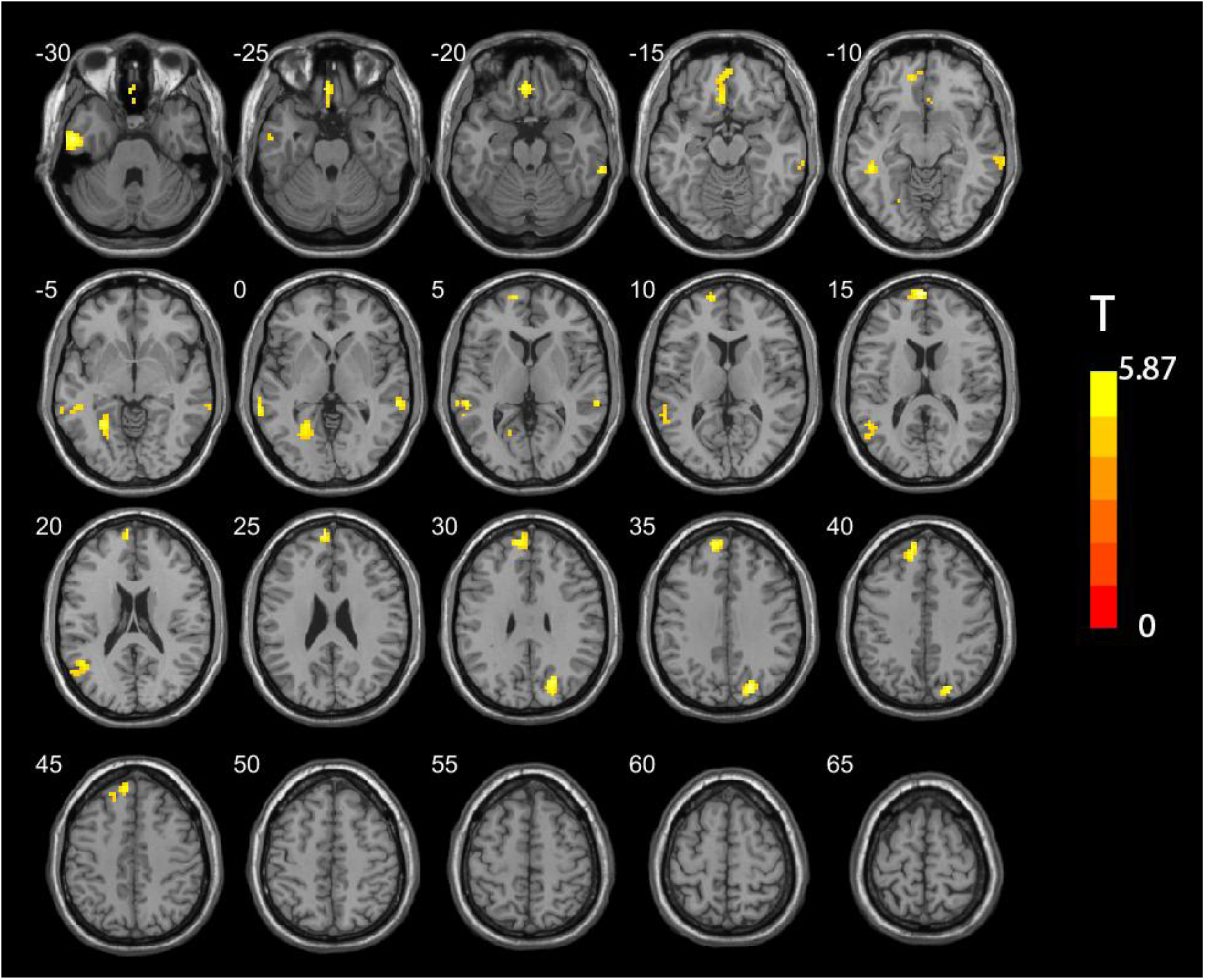
Altered functional connectivity between the left frontal lobe and other brain regions in IGE patients. The color bar represents the t-value. Warm colors indicate brain regions with significantly increased functional connectivity from the left frontal lobe in IGE patients compared with healthy controls. No regions with decreased functional connectivity were observed. The map threshold was set at voxel-level p < 0.001 and cluster-level p < 0.05, corrected by GRF.

**Fig. 4.**
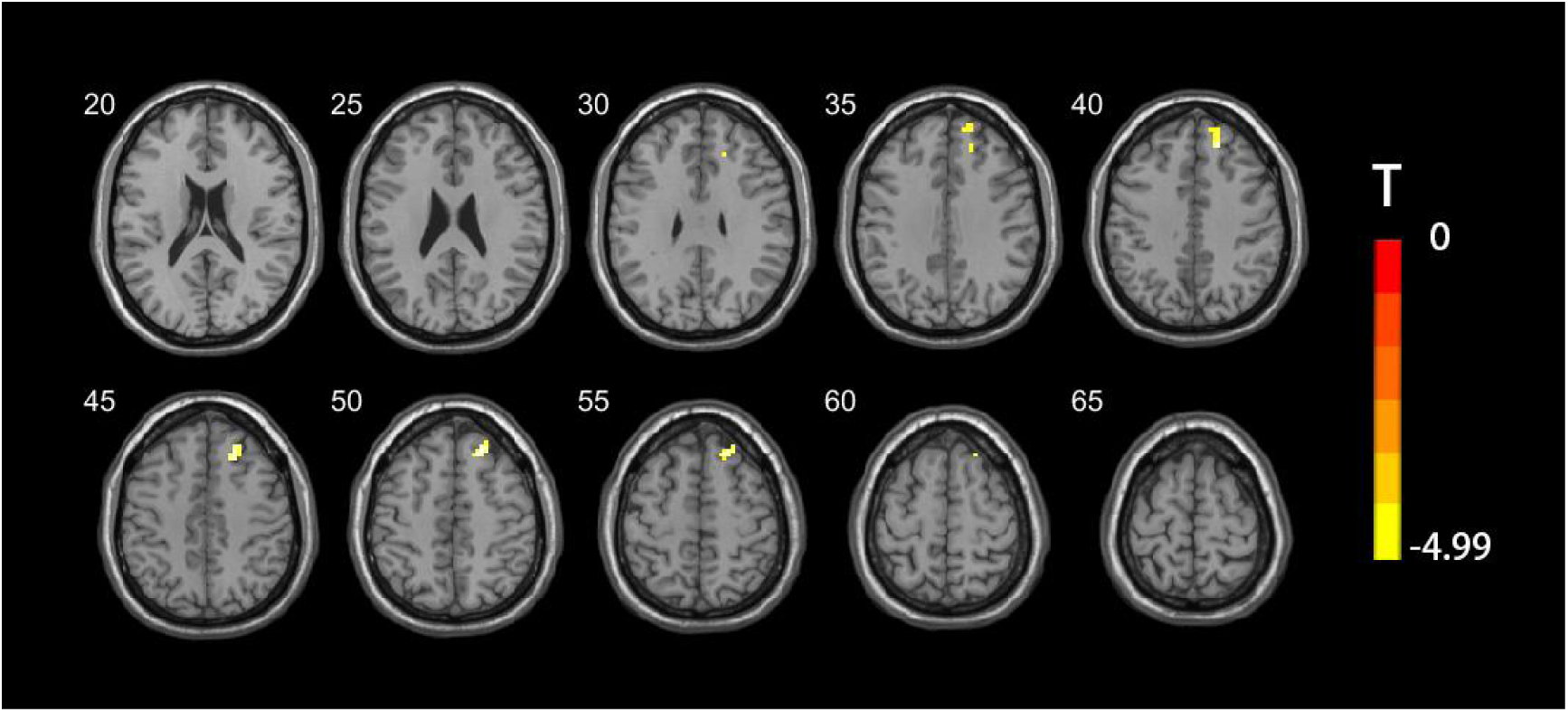
Altered functional connectivity between the right NAc and Frontal_Sup_2_R in the HMoCA group. The color bar represents the t-value. Compared with the LMoCA group, patients in the HMoCA group showed significantly decreased functional connectivity between the right NAc and Frontal_Sup_2_R. The map threshold was set at voxel-level p < 0.001 and cluster-level p < 0.05, corrected by GRF.

**Table 3.**
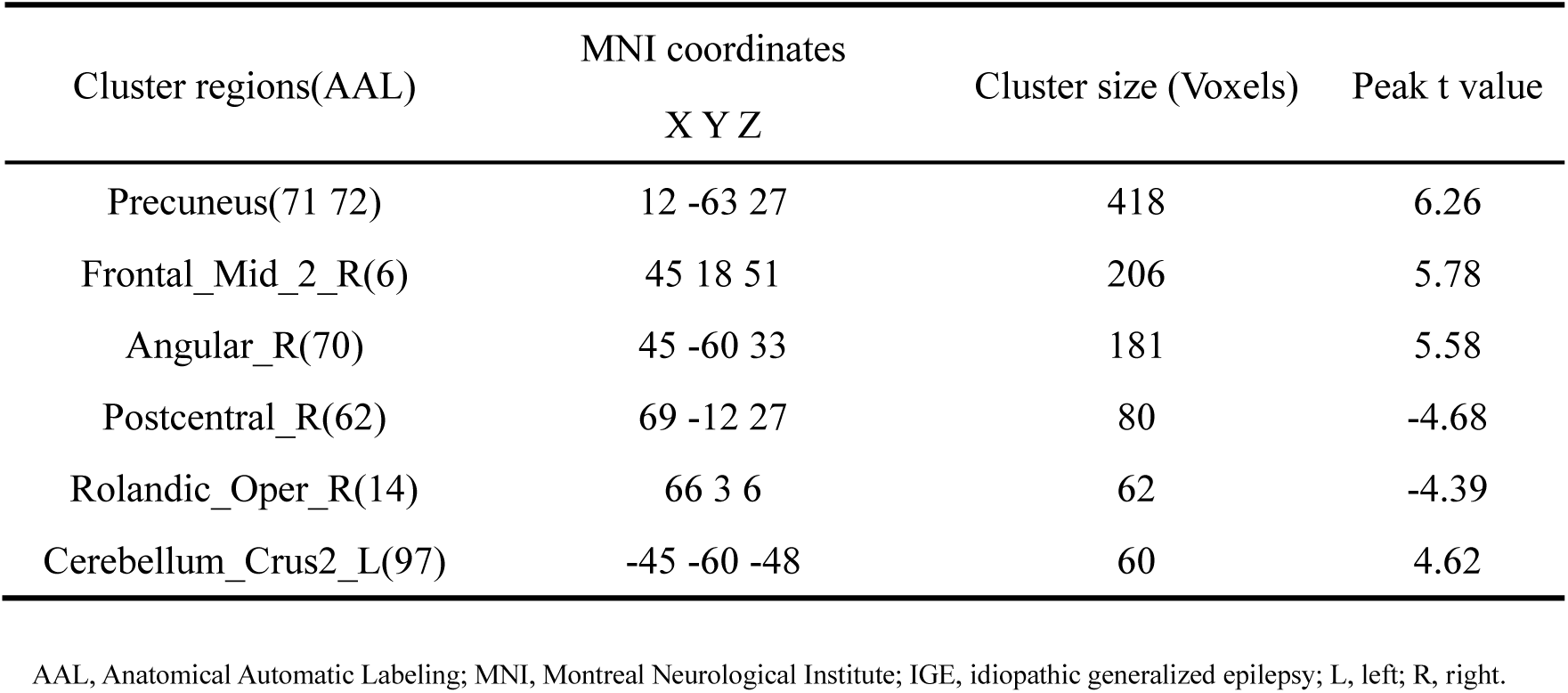
Altered FC from the right Cerebelum to other brain regions in IGE patients.

| Cluster regions(AAL) | MNI coordinates | Cluster size (Voxels) | Peak t value |
| --- | --- | --- | --- |
|  | X Y Z |  |  |
| Precuneus(71 72) | 12 -63 27 | 418 | 6.26 |
| Frontal_Mid_2_R(6) | 45 18 51 | 206 | 5.78 |
| Angular_R(70) | 45 -60 33 | 181 | 5.58 |
| Postcentral_R(62) | 69 -12 27 | 80 | -4.68 |
| Rolandic_Oper_R(14) | 66 3 6 | 62 | -4.39 |
| Cerebellum_Crus2_L(97) | -45 -60 -48 | 60 | 4.62 |

AAL, Anatomical Automatic Labeling; MNI, Montreal Neurological Institute; IGE, idiopathic generalized epilepsy; L, left; R, right.

**Table 4.** Altered FC from the left Frontal to other brain regions in IGE patients.

| Cluster regions(AAL) | MNI coordinates | Cluster size (Voxels) | Peak t value |
| --- | --- | --- | --- |
|  | X Y Z |  |  |
| Frontal_Sup_Medial_L(19) | -3 66 15 | 162 | 5.87 |
| Temporal_Mid_L(89) | -66 -42 -3 | 121 | 4.42 |
| Rectus_L(23) | -12 45 -12 | 120 | 4.76 |
| Lingual_L(51) | -27-54 -3 | 72 | 5.28 |
| Occipital_Sup_R(50) | 21 -75 33 | 69 | 5.14 |
| Temporal_Inf_L(93) | -57 -9 -30 | 58 | 4.90 |
| Temporal_Mid_R(90) | 63 -36 3 | 58 | 4.61 |

AAL, Anatomical Automatic Labeling; MNI, Montreal Neurological Institute; IGE, idiopathic generalized epilepsy; L, left; R, right.

**Table 5.** Altered FC from the right Nucleus Accumbens to other brain regions in HMoCA group.

| Cluster regions(AAL) | MNI coordinates | Cluster size (Voxels) | Peak t value |
| --- | --- | --- | --- |
|  | X Y Z |  |  |
| Frontal_Sup_2_R(4) | 21 33 51 | 72 | -4.98 |

AAL, Anatomical Automatic Labeling; MNI, Montreal Neurological Institute; IGE, idiopathic generalized epilepsy; L, left; R, right.

### Structural changes in JME and GTCSA subgroups of IGE patients with cognitive decline

We next studied if there are differences in neural signatures related to cognitive impairment between the two most common IGE subtypes [33–35]: Juvenile Myoclonic Epilepsy (JME) and Generalized Tonic-Clonic Seizures on Awakening (GTCSA). VBM analyses were performed to compare gray matter volumes between HMoCA and LMoCA groups within JME and GTCSA. In the JME group, patients with lower MoCA scores exhibited significantly increased GMV in the left angular gyrus, while in the GTCSA group, lower MoCA scores were associated with increased GMV in the left precentral gyrus. All findings survived GRF correction (voxel p < 0.005, cluster p < 0.05). These results suggest distinct structural correlates of cognitive impairment in different IGE subtypes.

In contrast, no significant differences were found in FC analyses or in correlation analyses within either subgroup. Full results are presented in Supplementary Table 6 and Supplementary Figure 1.

### Clinical characteristics associated with altered GMV and FC

We tested if the neuroimaging results would be sensitive enough to link to patients’ clinical data. Regarding GMV, we found that alterations in the right cerebellum, but not other areas (p>XX), were negatively correlated (r = -0.542, p = 0.001) with the course of the disease (Figure 5). For FC, no significant correlation (p>XX) with clinical data was found in IGE patients.

**Fig. 5.**
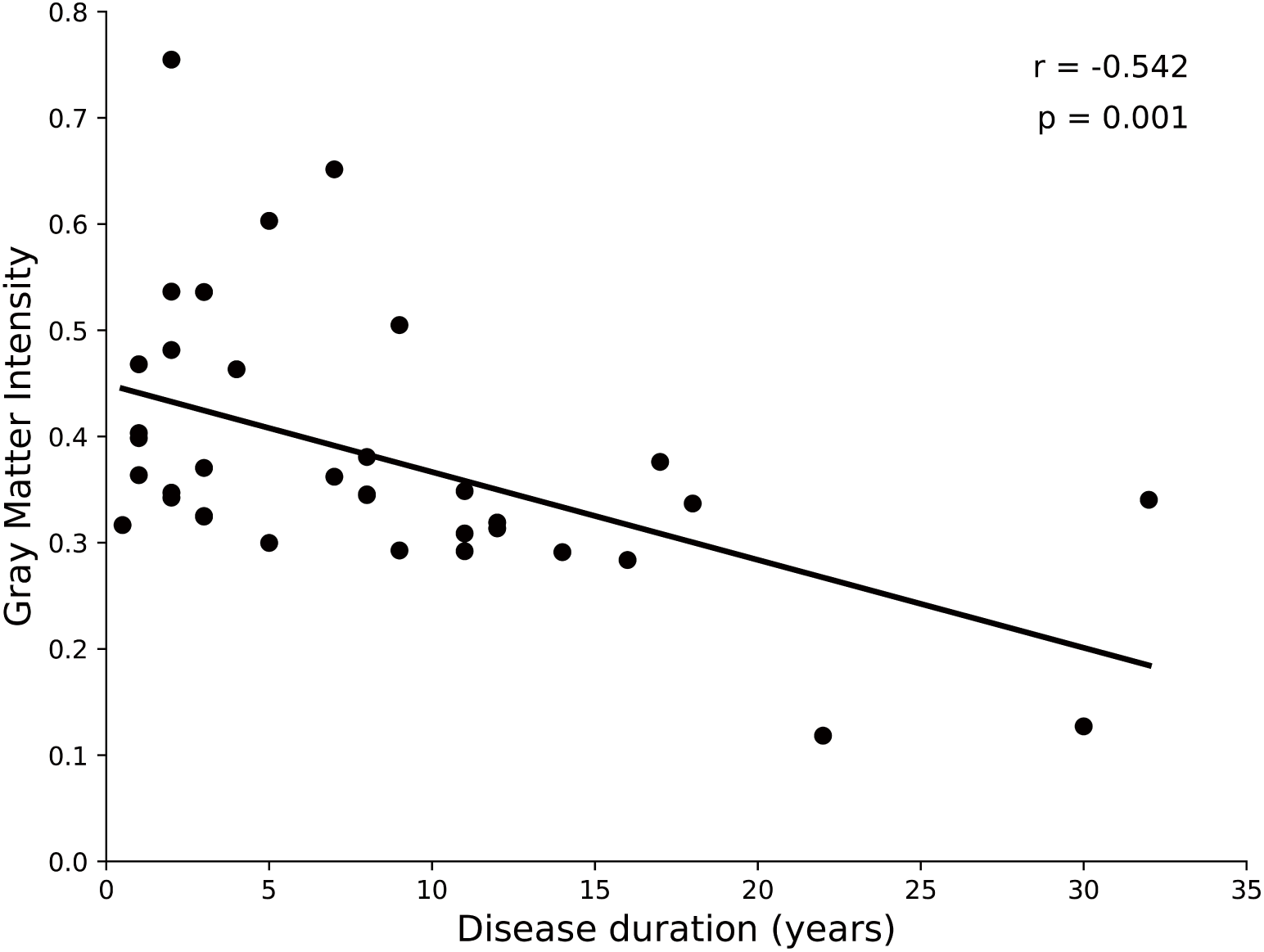
Correlation analysis between gray matter intensity in the right cerebellum and disease duration.

To further assess if there is any clinical characteristics related to brain signatures in cognitive impaired IGE patients, we developed various linear regression models. To assess the effects of GMV and FC on MoCA scores in the Cerebellum_8_R and Frontal_Sup_2_L regions. Univariate linear regression analyses of GMV and FC in these regions, after adjusting for sex, age, and years of education, showed no significant results (Supplementary Table 1). Multivariate linear regression analyses also failed to reveal significant associations (Supplementary Table 2). In sensitivity analyses, a substantial difference in FC for Frontal_Sup_2_L was observed in the univariate analysis (t = 2.13, *p* = 0.041) (Supplementary Table 3), but no significant results were found in the multivariate regression (Supplementary Table 4).

We also found significant differences in disease duration (t=2.72, p=0.010) and seizure frequency between the MoCA subgroups, therefore, we tested if the right NAc GMV and FC analyses remained statistically significant after controlling for these clinical variables. We found group differences in GMV (t = -4.413, p < 0.001 and FC (t = -2.683, p = 0.013) after controlling for variables (Supplementary Table 5), indicating the NAc structural and function effects we found are not caused by disease duration and seizure frequency. We also noted that the variance inflation factors (VIF) for all independent variables were below 2, indicating no significant multicollinearity issues in the model.

## Discussion

Using VBM with functional connectivity analysis, this study comprehensively examined the structural and functional brain connectivity changes in patients with IGE. We here revealed several significant findings that may be closely related to the neuropathological mechanisms of IGE and provided new insights for further understanding of cognitive dysfunction in IGE.

MoCA is a comprehensive assessment tool for cognitive functioning that measures several aspects of cognitive ability, such as memory, language, attention, and execution, reflect the cognitive differentiation of IGE patients[36, 37]. Previous studies have shown that NAc plays a crucial role in cognitive tasks such as executive function, cognitive flexibility, and reward processing[19, 38]. A recent research also suggested that the nucleus accumbens is closely associated with seizures in animal models of epilepsy and can exert antiepileptic effects by precisely modulating GABAergic neurons[39], which is supported by Zhao et al. [40] showing that the shell portion of the nucleus accumbens in human patients with temporal lobe epilepsy (TLE) exhibits significant impairment of fiber integrity and connectivity abnormalities, forming part of the characteristic changes of the seizure part of the characteristic changes in the network. In this study, we surprisingly found an increased GMV in the right NAc. Similar to previous research that showed a compensatory effect in cognitively impaired patients[41], our results showing an increased NAc GMV, which could be linked to a specific compensatory effect for cognitive deficits in IGE patients.

We also observed an increased FC between the right NAc and superior frontal gyrus in patients in the LMoCA group. While NAc forms an essential neural network with the prefrontal cortex, amygdala, and other regions to regulate emotional responses, motivated behaviors, and executive functions [42, 43]. We speculate that enhanced connectivity between the nucleus accumbens and the superior frontal gyrus may, to some extent, compensate for cognitive deficits in patients with IGE [44], which could serve as a potential biomarker for early identification of cognitive decline in IGE patients.

Notably, MoCA-based subgroup analyses revealed divergent neuroanatomical patterns between the overall IGE cohort and its two major subtypes, JME and GTCSA [4, 45–47]. In the overall cohort, lower MoCA scores were associated with increased GMV in the right NAc and heightened FC with prefrontal regions, suggesting a broader network-level response. In contrast, the JME and GTCSA subtypes exhibited increased GMV in the left angular and precentral gyri without corresponding FC alterations. These differences may indicate distinct mechanisms of cognitive impairment across the IGE spectrum. The localized frontal GMV increases in JME and GTCSA could reflect subtype-specific cortical reorganization, whereas the accumbens-prefrontal changes observed in the overall cohort may represent a more generalized compensatory or maladaptive response. Given the relatively small sample sizes in the subtype analyses, especially for FC, these findings should be interpreted with caution. Nevertheless, the structural alterations identified were statistically robust and anatomically distinct, supporting their potential biological relevance.

Further research with larger, subtype-stratified cohorts is warranted to validate these findings and elucidate the underlying mechanisms of cognitive dysfunction in IGE.

Our evidence showed that IGE patients has reduced right cerebellar GMV in comparison with healthy controls, and that the reduction is negatively correlated with disease duration. This result supports previous studies on structural changes in the cerebellum in patients with epilepsy, especially its importance in cognitive and emotional functions [48–50]. Recent studies have shown that the cerebellum is involved in motor control and plays a role in cognitive regulation and emotional control[51]. Cerebellar functions show lateralization, with the left cerebellum more often associated with spatial abilities, while the right cerebellum is mainly associated with language and executive functions [52–54]. Moreover, the presence of separated neural loops between the cerebellum and frontal lobe has been found to support the idea that the cerebellum has different functional subdivisions in motor and cognitive control [55]. The GMV decrease in the right cerebellar area VIII observed in the present study in IGE patients increased with the duration of the disease, suggesting cumulative damage to the cerebellum from prolonged seizures. Such structural changes may affect the cerebellar structure through mechanisms such as neuronal damage and altered neuroplasticity and may also lead to diminished neural network functioning, which may trigger impairments in cognition and emotion regulation in patients with epilepsy [56]. In addition, long-term antiepileptic drug treatment in patients with IGE may contribute to cerebellar volume changes, potentially through neuroprotective mechanisms, effects on neuronal metabolism, or drug-induced neuroplasticity [57, 58]. Future research could further focus on tracking unmedicated early IGE patients through longitudinal designs. This is to exclude drug interference with cerebellar structures and to deeply explore the dynamic effects of epilepsy itself on cerebellar structure and functional networks, thereby providing new targets for early disease intervention.

As part of the prefrontal cortex, the left dorsolateral superior frontal gyrus plays an important role in executive function, cognitive control, and attentional regulation [59]. Previous studies indicate that epilepsy patients may exhibit enhanced function in prefrontal regions, potentially driven by neuroplastic changes that compensate for the effects of seizures on other brain areas [60, 61]. The increased GMV we observed may reflect this adaptive change to compensate for the effects of cerebellar structural changes on cognitive functioning [62]. Significantly, the increase in the left superior frontal gyrus may also be due to the patient’s increased demand for cognitive resources, which leads to an increase in the volume of grey matter in this region [63].

We also found various FC altered with the rs-fMRI in IGE patients. These include FC abnormalities linked to the precuneus and angular gyrus, which are associated with visuospatial attention and language processing [64], as well as FC abnormalities linked to frontal regions, related to executive functions and cognitive control [65]. The enhanced cerebellar connectivity to these brain regions may be an adaptive response to cognitive impairment in IGE patients due to epilepsy. Reduced FC of the right cerebellum to the postcentral gyrus and Rolandic operculum, on the other hand, may reflect underlying impairments in sensory and motor control in patients with IGE [15, 66].

## Limitation

While our study provides valuable insights into the altered brain structure and functional connectivity underlying cognitive impairment in IGE, several limitations should be considered when interpreting the findings. First, the measurement tools used in this study may have technical limitations that hindered the accurate detection of subtle changes in brain regions or functional connectivity. fMRI offers advantages in spatial resolution, providing detailed information about brain networks. However, compared to electroencephalography (EEG) and magnetoencephalography (MEG), fMRI has limitations in temporal resolution. The temporal resolution of fMRI is constrained by hemodynamic responses, which occur on a timescale of seconds, making it less suitable for capturing rapid neural dynamics detected with millisecond precision by EEG/MEG. However, fMRI is particularly valuable for studying connections in deep brain structures, such as the cerebellum, where EEG/MEG has limited sensitivity [67].

Second, the small sample size in this study may have led to insufficient statistical power, preventing the detection of potentially significant relationships. Increasing the sample size would improve the reliability of the findings, and future multi-centre studies will enhance the comprehensiveness of the experimental design. Third, this study did not perform subgroup analyses for drug-naïve or newly diagnosed epilepsy patients, which limits our ability to exclude potential medication effects. There may also be uncontrolled confounding factors that affect the relationship between GMV, FC, and MoCA scores. The genetic basis of IGE is important, and future research could further incorporate family history information and consider genetic analyses to study the impact of genetics on brain structure and cognitive function in IGE patients.

While a quick and easy screening tool, MoCA provides only a preliminary assessment of overall cognitive function. However, it does not offer a detailed evaluation of specific cognitive domains, such as executive function, memory, and attention, which is a limitation of this study. It is important to note that this study’s observed structural/functional connectivity between MoCA score subgroups does not imply causality. As a cross-sectional study, participants were only scanned once, and longitudinal changes in disease progression could not be assessed. The directionality of these changes remains unclear, and it is uncertain whether the changes in connectivity cause or consequence of cognitive impairment. To address this, techniques like Dynamic Causal Modeling (DCM) can reveal the direction of information flow and provide a more comprehensive understanding of brain network dynamics[68]. Finally, we acknowledge the clinical heterogeneity within the IGE cohort, including variability in disease duration, seizure exposure, and subtype differences. Our findings suggest that some brain alterations are associated with cognition independently of clinical variables, whereas others may be influenced by disease chronicity. In the future, we aim to recruit a larger and more diverse cohort and apply a longitudinal design with detailed neuropsychological testing to address these limitations and validate the current findings.

## Conclusion

This study revealed abnormalities in brain structural and functional connectivity in patients with IGE and explored the relationship between cognitive function and brain imaging features through MoCA stratification analysis. These findings suggest that changes in structural and functional connectivity may serve as potential neurobiological markers of IGE, providing new insights into the neuropathological mechanisms of epilepsy. Future studies could further examine the long-term effects of specific changes in these brain regions on cognitive function, especially in unmedicated or early-stage patients, to help clarify the direct effects of epilepsy on the brain and its potential impact on cognitive function.

## Acknowledgement

This work was supported by the National Natural Science Foundation of China [Grant Number 8207145]. The authors thank all participants and their families for their involvement in this study. We are also grateful to the clinicians and research staff for their assistance in data collection.

