## Supplementary Figure 1 for "Structural and functional changes linked to cognitive impairment in Idiopathic Generalized Epilepsy"

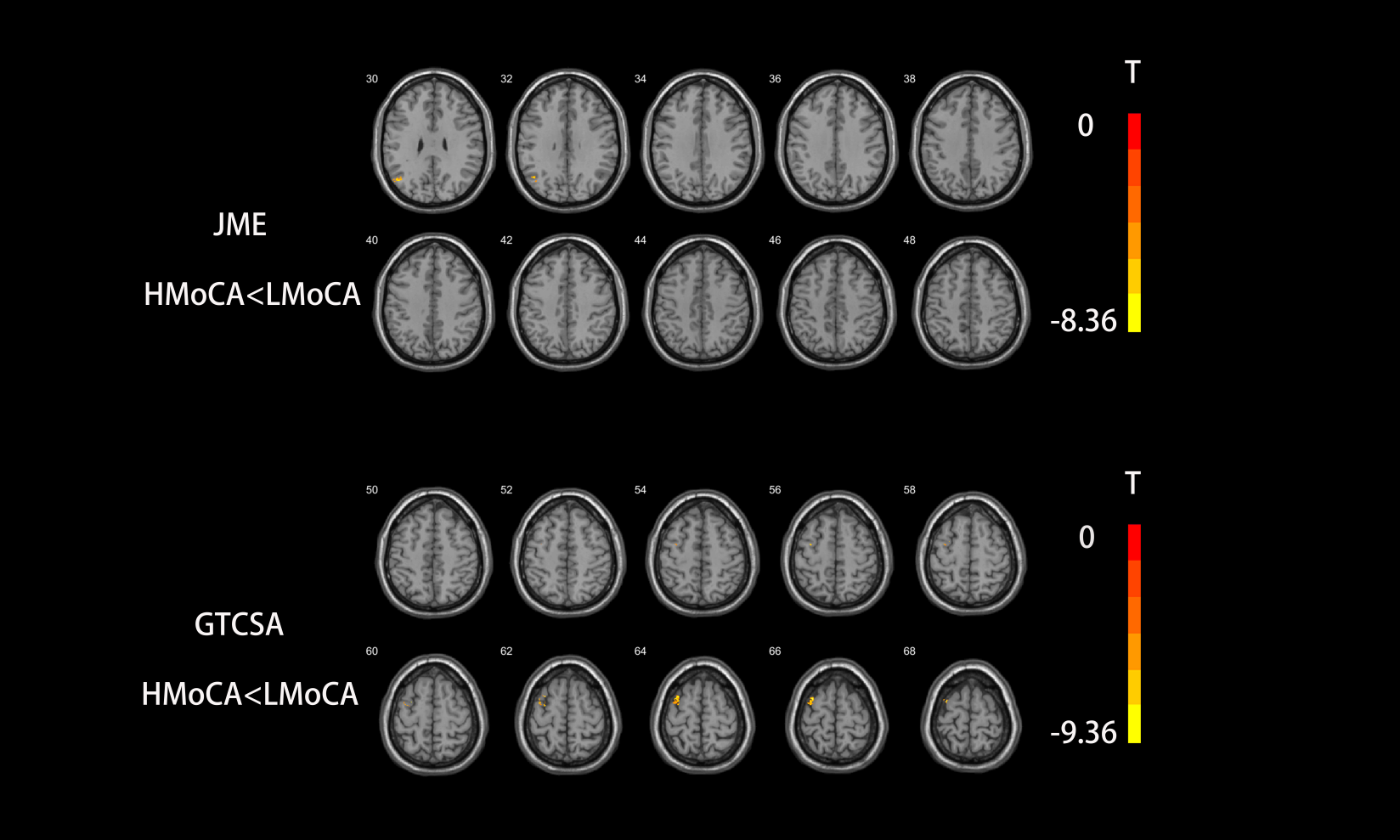

Supplementary Fig. 1 Voxel-based morphometry results showing gray matter volume differences between high and low MoCA groups in patients with juvenile myoclonic epilepsy (JME, top) and generalized tonic–clonic seizures alone (GTCSA, bottom). Statistical comparison was performed using a two-sample t-test. The threshold was set at p < 0.05 (GRF-corrected). The color bars represent the T-values.
