## Supplementary Table for "Structural and functional changes linked to cognitive impairment in Idiopathic Generalized Epilepsy"

Supplementary Table 1: Linear regression of MoCA scores based on GMV and FC in significant brain regions: Cerebellum_8_R and Frontal_Sup_2_L, adjusted for gender, age, and years of education.

|  | B | SE | t | *p* value |
| --- | --- | --- | --- | --- |
| Cerebelum_8_R GMV | -0.26 | 0.41 | -0.65 | 0.523 |
| Cerebelum_8_R FC | 0.58 | 0.40 | 1.46 | 0.155 |
| Frontal_Sup_2_L GMV | 0.07 | 0.41 | 0.18 | 0.858 |
| Frontal_Sup_2_L FC | 0.57 | 0.40 | 1.43 | 0.162 |

MoCA, Montreal Cognitive Assessment; IGE, Idiopathic Generalized Epilepsy; GMV, gray matter volume; FC, functional connectivity; SE, Standard Error.

Supplementary Table 2 Multiple linear regression of MoCA scores based on GMV and FC in significant brain regions: Cerebelum_8_R and Frontal_Sup_2_L, adjusted for gender, age, and years of education.

|  | B | SE | t | *p* value | VIF |
| --- | --- | --- | --- | --- | --- |
| Cerebelum_8_R GMV | -0.16 | 0.42 | -0.37 | 0.713 | 1.05 |
| Cerebelum_8_R FC | 0.43 | 0.43 | 1.00 | 0.325 | 1.13 |
| Frontal_Sup_2_L GMV | 0.23 | 0.43 | 0.54 | 0.596 | 1.13 |
| Frontal_Sup_2_L FC | 0.47 | 0.46 | 1.03 | 0.312 | 1.27 |

MoCA, Montreal Cognitive Assessment; IGE, Idiopathic Generalized Epilepsy; GMV, gray matter volume; FC, functional connectivity; SE, Standard Error; VIF, Variance Inflation Factor.

Supplementary Table 3: Linear regression of MoCA scores based on GMV and FC in significant brain regions (Cerebellum_8_R and Frontal_Sup_2_L), adjusted for gender, age, education, ASMs count, seizure frequency, and disease duration.

|  | B | SE | t | *p* value |
| --- | --- | --- | --- | --- |
| Cerebelum_8_R GMV | -0.28 | 0.39 | -0.72 | 0.474 |
| Cerebelum_8_R FC | 0.48 | 0.39 | 1.25 | 0.219 |
| Frontal_Sup_2_L GMV | -0.05 | 0.40 | -0.11 | 0.910 |
| Frontal_Sup_2_L FC | 0.80 | 0.38 | 2.13 | 0.041^*^ |

MoCA, Montreal Cognitive Assessment; IGE, Idiopathic Generalized Epilepsy; GMV, gray matter volume; FC, functional connectivity; ASMs: anti-seizure medications; SE, Standard Error. ^*^, *p*＜0.05.

Supplementary Table 4 Multiple linear regression of MoCA scores based on GMV and FC in significant brain regions (Cerebellum_8_R and Frontal_Sup_2_L), adjusted for gender, age, education, ASM count, seizure frequency, and disease duration.

|  | B | SE | t | *p* value | VIF |
| --- | --- | --- | --- | --- | --- |
| Cerebelum_8_R GMV | -0.15 | 0.39 | -0.39 | 0.700 | 1.05 |
| Cerebelum_8_R FC | 0.22 | 0.41 | 0.53 | 0.602 | 1.18 |
| Frontal_Sup_2_L GMV | 0.16 | 0.41 | 0.40 | 0.691 | 1.13 |
| Frontal_Sup_2_L FC | 0.75 | 0.45 | 1.68 | 0.104 | 1.36 |

MoCA, Montreal Cognitive Assessment; IGE, Idiopathic Generalized Epilepsy; GMV, gray matter volume; FC, functional connectivity; SE, Standard Error; ASMs: anti-seizure medications; VIF, Variance Inflation Factor.

Supplementary Table 5: Regression Coefficients of MoCA Group Predicting Right Nucleus Accumbens (N_Acc_R) Gray Matter Volume (GMV) and Functional Connectivity (FC)

| Outcome | B | SE | t | *p* value | VIF |
| --- | --- | --- | --- | --- | --- |
| N_Acc_R GMV | -0.071 | 0.016 | -4.413 | 0.000^*^ | 1.528 |
| N_Acc_R FC | -0.078 | 0.029 | -2.683 | 0.013^*^ | 1.528 |

Multiple linear regression models were conducted to assess the effect of MoCA subgroup (HMoCA = 1, LMoCA = 0) on right nucleus accumbens GMV and FC, controlling for age, sex, years of education, total intracranial volume (TIV), number of antiseizure medications (ASMs), Idiopathic Generalized Epilepsy subtypes, disease duration, and seizure frequency. Only the regression coefficients for the MoCA group are reported here. MoCA, Montreal Cognitive Assessment; SE, Standard Error; VIF, Variance Inflation Factor. ^*^, *p*＜0.05.

Supplementary Table 6: Results of VBM analyses between high and low MoCA groups in patients with JME and GTCSA.

| Groups | Cluster regions(AAL) | MNI coordinates | Cluster size (Voxels) | Peak t value | p value |
| --- | --- | --- | --- | --- | --- |
|  |  | X, Y, Z |  |  |  |
| JME  HMoCA＜LMoCA | Angular_L (65) | -43.75 -62.25 30.75 | 110 | -8.36 | 0.00^**^ |
| GTCSA  HMoCA＜LMoCA | Precentral_L (1) | -30.75 3.75 68.75 | 362 | -9.36 | 0.00^**^ |

AAL, Anatomical Automatic Labeling; MNI, Montreal Neurological Institute; JME, juvenile myoclonic epilepsy; GTCSA, generalized tonic–clonic seizures alone; L, left; R, right. ^*^ p＜0.05; ^**^ p＜0.01.
